# Understanding SARS-CoV-2 Infection and Dynamics with Long Term Wastewater based Epidemiological Surveillance

**DOI:** 10.1101/2021.03.15.21253574

**Authors:** Athmakuri Tharak, Harishankar Kopperi, Manupati Hemalatha, Uday Kiran, C. G. Gokulan, Shivranjani Moharir, Rakesh K Mishra, S Venkata Mohan

## Abstract

Wastewater-based epidemiology (WBE) of SARS-CoV-2 emerged as an advantageous method to study the infection dynamics at substantial population level. A temporal glimpse at sewage viral genome helps as diagnostic tool to understand the viral spread at community level. In this study, for the long-term epidemiological surveillance, we monitored the SARS-CoV-2 genetic material in domestic sewage by adopting the longitudinal sampling to represent a selected community (∼1.8 lakhs population which occupies 1.79% of the total population of Hyderabad city) to understand the dynamics of infection. Dynamics and spread of COVID-19 outbreak within the selected community were achieved by studying the longitudinal sampling for a specific period of time. WBE also promotes clinical scrutiny along with disease detection and management, in contrast to an advance warning signal to anticipate outbreaks.

## 1. Introduction

The diagnostic aids, equipment and facilities were phenomenally improved since the pandemic of SARS-Cov-2 emergence, besides the surveillance of SARS-CoV-2 by clinical data employing swab samples from person to person in critical periods of the pandemic is a challenge to understand spread among the communities (Asgar et al., 2014; O’Reilly et al., 2020). Wastewater based epidemiological study (WBE) is being conceived as one of the standard protocols that help to infer the dynamics and infection of the SARS-CoV-2 and its state of severity among the community (Xagoraraki and O’Brien, 2020; Shaw et al., 2020; Randazoo et al., 2020; Venkata Mohan et al., 2021). WBE data will assist new viruses to be detected in a community previous to clinical recognition that allows impart the preventive measure and precautions among the community to resist the outbreak (Casanova et al., 2015; Brainard et al., 2017; Torrey et al., 2019). The ability of SARS-CoV-2 to infect the gastrointestinal tract (GI) in addition to the bronchial inflammation is well reported (Ahmed et al., 2020a; Wu et al., 2020; Ahmed et al., 2020b; Wurtzer et al., 2020; La Rosa et al., 2020; Medema et al., 2020; Usman et al., 2020). The design of the sampling protocol is a crucial factor to detect the COVID-19 genetic material in the wastewater. A load of viral material in sewage alters temporally based on the time of defecation frequencies and sampling (Weidhaas et al., 2020; Nakamura et al., 2015). Diverse viral load shedding from the affected community, converging of household wastewater and industrial effluents and time of sampling could affect the detection of the viral genome in the sewage, however, WBE provides a range of information to predict the dynamics of infection with the design of sampling protocol (Ahmed et al., 2020; Wurtzer et al., 2020; Venugopal et al., 2020; Kopperi et al., 2021; Daughton et al., 2018; Lednicky et al., 2020; Venkata Mohan et al., 2021; Quilliam et al., 2020). Different independent SARS-CoV-2 WBE studies followed different water sampling and processing methods to detect the SARS-CoV-2 RNA in sewage (Daughton 2018; Lednicky et al., 2020; Venkata Mohan et al., 2021; Quilliam et al., 2020), The grab sampling method can be used in remote areas and even in poor sewage system conditions, which helps in better surveillance (Kopperi et al., 2021). These WBE methods will help to monitor surveillance in the agency area where proper infrastructure and hospitality lacks. A true sampling at the selected station and data obtaining from the study provides information about the spread and impact of the severity that helps to give the alarming signal to the corresponding community (Mao et al., 2020; Hart et al., 2020; Hemalatha et al., 2021).

In the present study, an attempt was made to investigate the persistence and dynamics of SARS-CoV-2 in domestic sewage by conducting longitudinal sampling over a period of six months (July 2020 to February 2021 excluding the rainfall event months i.e., August 2020 and September 2020) in a selected community representing 1.8 lakhs of population.

## 2. Methodology

### 2.1 Sampling area

The study area represents a community with ∼1.8 lakhs population covering Tarnaka, HMT Nagar, Lalaguda and Nacharam as part of Greater Hyderabad, Telangana (State), India. The selected community discharges 18 MLD of domestic wastewater (sewage) flows through the main drain starting from Lalaguda finally covering at sampling point T10 before STP inlet (Fig 1). Various laterals drain joins the main drain covering the adjoining domestic settlements. Eight sampling points were selected across the drain system to comprehensively represent the majority of lateral drains (Fig. 1). Sampling points were selected in such a way to cover the entire community sewage network. Samples were collected at the lateral drain before getting merged into the main drain. The main sewage drain of the community finally gets discharged into the sewage treatment plant (STP; 10 MLD) located at Nacharam.

**Fig. 1.**
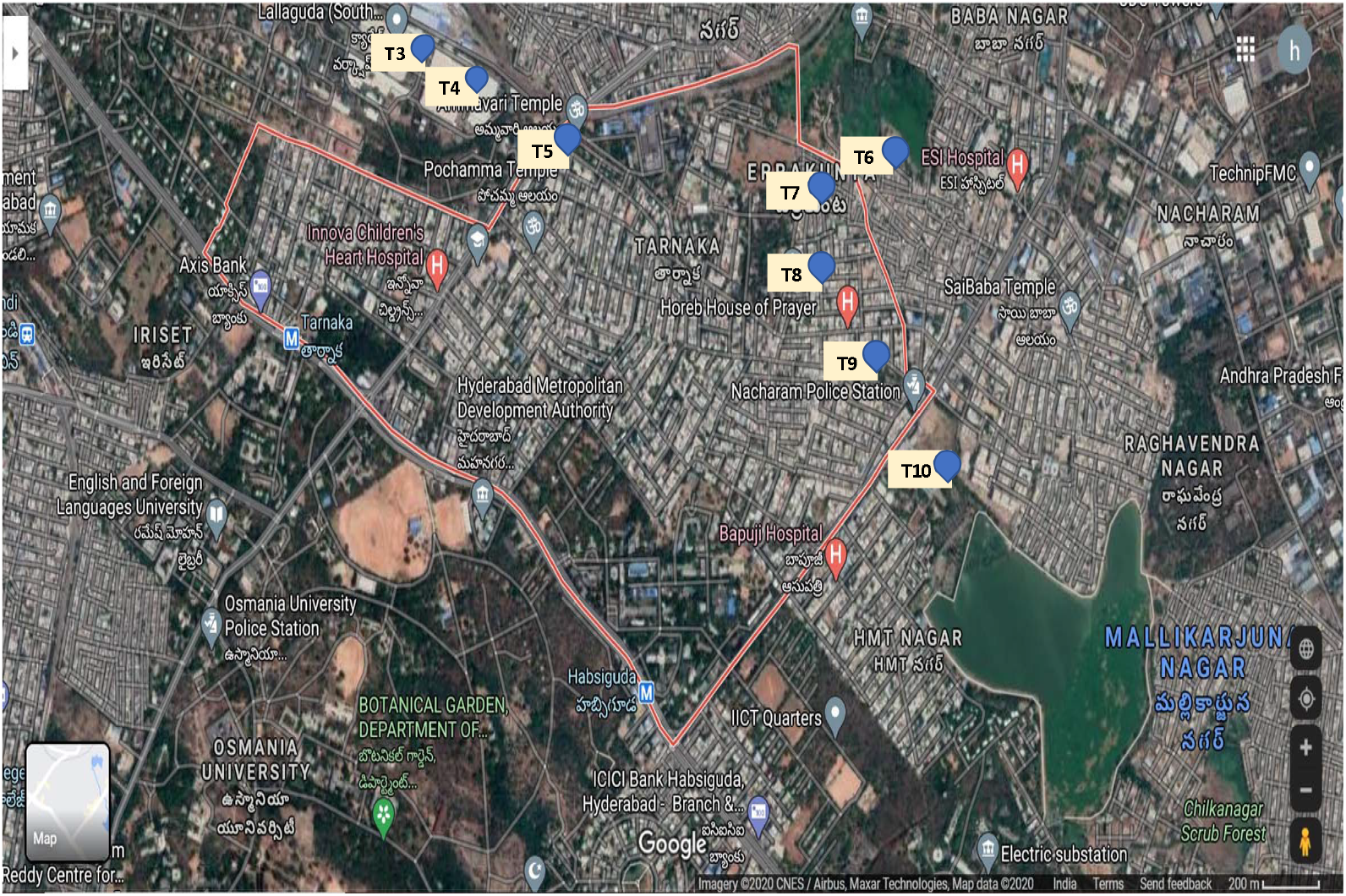
Map showing the point of sample collection (Tarnaka and Nacharam) (Courtesy: Google Map)

### 2.2 Details of Sampling

Grab sampling protocol was followed to sample domestic wastewater at the selected sampling stations (APHA 2017). Samples were collected at 8:00 to 8:30 am on the day wherein there was no rainfall event, 2 days prior to sampling day also. Grab samples were collected on a weekly and monthly basis. Total of eight samples were collected for weekly monitoring starting from 07-10-2020 (Week 1), 28-10-2020 (Week 4), 04-11-2020 (Week 5), 11-11-2020 (Week 6) and 18-11-2020 (week 7). Samples were not collected during Week 2 and Week 3 due to heavy rainfall events that occurred leading to the overflow of all sewage drains. Monthly samples were sampled at terminal covering point of the main drain (T10) starting from July 2020 and planned to collect over the 6 months continuously, but due to the monsoon rainfall in early august (14/08/2020 to 24/09/2020 and 10/10/2020 to 21/10/2020), sampling was paused and restarted from October 2020 continued till March 2021 (Table 1).

**Table 1:**
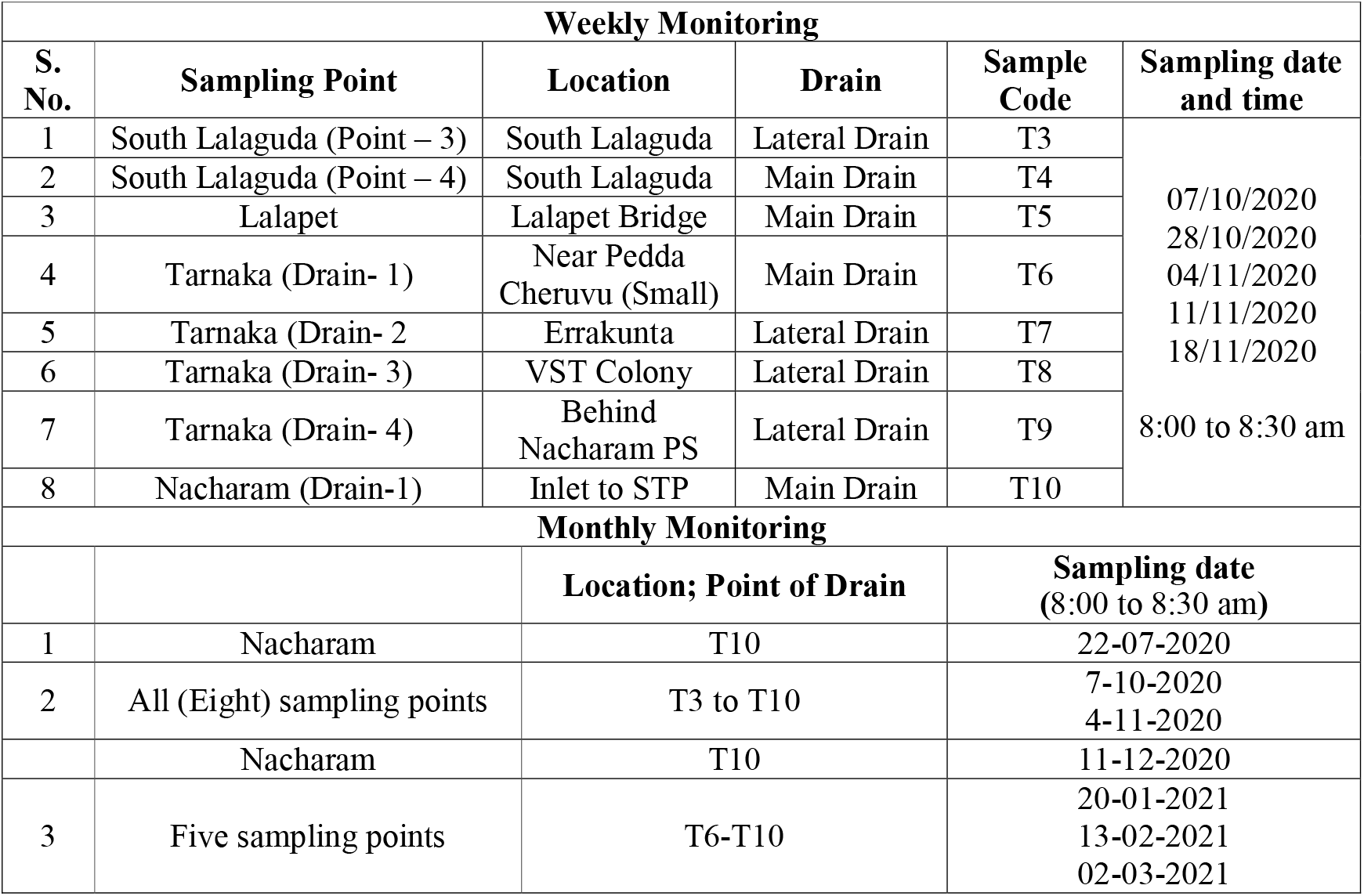
Details of Sampling with reference to time of weekly and monthly samples (As shown in Fig 1)

### 2.3 Sample collection and processing

Samples were collected with all the safety measures as discussed in Hemalatha et al., 2021. The sample container was slightly lowered in the opposite direction of flow with partial immersion. After sampling the external surface of the container is disinfected with 70% ethanol to prevent contamination and sealed with plastic bags, labeled and transported (4±1 °C) immediately to the lab and stored at 4°C until further processing. Samples were processed within 12 h.

### 2.4 Processing of Samples

Collected samples were subjected to the gravity filtration using 1 mm filter papers to remove the larger debris followed by secondary filtration with 0.2 µm filtration units (Nalgene® vacuum filtration system) to remove other fine particles and pathogens (Hemalatha et al., 2021). 60 mL of the total filtrate was concentrated to ∼600 µl using 15 mL 30 kDa Amicon® Ultra-15 (Merck Millipore) by ultra-filtration (4000 rpm; 4 °C; 10 min). 150 μL of the concentrated sample was used for RNA extraction. All the sample processing and detection experiments were performed in a Biosafety level 2 (BSL-2) laboratories.

### 2.5 RNA extraction and RT-PCR

RNA was extracted from the concentrated samples using the Viral RNA isolation kit (QIAamp, Qiagen) with provided manufactures protocol. DNA/RNA cross-contamination was avoided by using sterile equipment and RNase-free water for the RNA extraction (Hemalatha et al., 2021; Kopperi et al., 2021). Isolated SARS-CoV-2 RNA was quantified by using FDA (Food and Drug Administration, USA Government) approved RT-PCR Detection Kit (Shanghai Fosun Long March Medical Science Co., Ltd, China). Fosun RT-PCR containing the primers and chromophore probes encoding for the envelope protein-coding gene (E-gene; ROX), nucleocapsid gene (N-gene; JOE), and open reading frame1ab (ORF1ab; FAM) of SARS-CoV-2. RT-PCR reaction procedure includes two initial cycles, Reverse transcription (50°C for 15 min) and the Initial denaturation (95°C for 3 minutes) followed by 45 cycles at 95°C for 5 seconds and 60°C for 40 seconds. Signals from the probes (FAM (ORF1ab), JOE (N gene), ROX (E gene), and CY5 (Internal reference)) were collected by the fluorescence channels at 60°C. Both positive and negative controls of the fosun RT-PCR kit were included in all the amplifications. C_T_ values in the positive controls match with given manufactured data and no C_T_ was observed in negative control that states the devoid of contamination. All the samples done were analyzed in triplicate.

### 2.6 Statistical methods and data management

To calculate the number of RNA copies per litre of collected domestic wastewater samples, linear fit equation of the E-gene was employed and RNA copies per liter wastewater was calculated (Hemalatha et al., 2021).

## 3. Results and Discussion

### 3.1 Weekly Sample Analysis for SARS-CoV-2

SARS-CoV-2 genetic material was detected in all the 40 samples collected at 8 sampling stations over the window period of five weeks from 07/10/2020 to 18/11/2020 with variable loads. Amplification of three SARS-CoV-2 target genes, namely, E gene, N gene, and ORF1ab was detected in all the samples. Apart from the C_T_ values, to predict the SARS-CoV-2 viral load in domestic sewage, the RNA copy number was calculated considering the linear fit drawn from the standard curve of the E-gene, in our previous work (Hemalatha et al., 2021). C_T_ values (average) of E-gene, N-gene, and ORF1ab at the initial sampling point (South Lalaguda lateral drain; T3) were 28.35±1.27%, 26.85±1.30%, and 27.89±0%, respectively with five-week average RNA copies of 23,470 Copies/L. The extended point to the T3, that is the T4 sample (South Lalaguda Main drain) showed C_T_ values (average) of 27.16±1.29%, 25.39±1.65%, and 21.01±0.32% for E-gene, N-gene, and ORF1ab, respectively with the RNA copy number of 54,135 Copies/L. An increase in the RNA copy number at the T4 might be due to the converging of the lateral drains containing domestic sewage discharge into the main drain. Wastewater from the main drain (T4) flows continuously till the end of the selected longitudinal sampling point (Nacharam Inlet to STP; T10). The third sampling point located at Lalapet Bridge (T5) showed relatively less C_T_ values (Average; E-gene, 28.73±1.04%; N-gene, 27.54±1.82%; ORF1ab were, 28.72±0.93%) compared to T3 and T4 corresponding to 17,954 RNA Copies/L. Even though the main drain stream continued from the earlier sampling point, the number of RNA copies was reported less at the T5 that might be due to the conflate of diary processing effluents with excessive surfactants (chemical) discharged into the drain that may disintegrate the viral RNA material. Similar observations were reported elsewhere (Yonar et al., 2018; Barcelo 2020; Westhaus et al., 2021). Downstream T5, the domestic sewage overflow from Pedda Cheruvu (Lake) and discharges into the main drain (T6). C_T_ values (average) at T6 was recorded to be 27.42±3.36%, 26.28±4.68%, and 27.14±0% for E-gene, N-gene, and ORF1ab, respectively with RNA copies of 45050 Copies/L. Relative increment in viral load was evident with T6 samples compared to T5. In addition to samples collected in the main drain, we have collected samples of lateral drains that were flowing from the set of communities located around the main drain and this flow finally merges into the main drain. These lateral drain sewage samples depicted marginally low values of SARS-CoV-2 load than the main drain.

C_T_ values (average) of E-gene, N-gene, and ORF1ab of the samples collected at T7 (Errakunta lateral drain) was 28.46±2.39%, 26.58±2.67%, and 21.61±1.31% respectively with 21757 Copies/L. Similarly, the two lateral drains T8 (VST lateral drain) and T9 (Nacharam lateral drain) recorded C_T_ values (average) of 26.03±1.96/25.41±1.50% (E-gene), 24.53±3.38%/23.37±3.34% (N-gene) and 20.06±1.21%/19.56±1.33% (ORF1ab) with higher RNA copies 119391 and 184664 RNA Copies/L, respectively. T10 samples representing the terminal point of main drain where the flow of all the previous sampling points converges showed C_T_ values that was almost an average of those obtained from other sampling sites individually (C_T_ values (average): 27.24±1.84% (E-gene), 25.71±1.28% (N-gene), 21.33±0.48% (ORF1ab)) with 51182 RNA Copies/L. C_T_ values of E-gene in all the sampling points range between 25.41±1.50% and 28.73±1.04% with an average of 27.35±1.83% (Table 2). Similarly, the C_T_ values of the N-gene and ORF1ab was observed between 23.37±3.34% to 27.54±1.82% and 19.56±1.33% to 28.72±0.93% respectively having an average of 25.78±2.48%, and 23.42±0.7% (Table 2). Along with the cumulative average, individual three genes average value of sampling points from T3 to T9 correlated with the C_T_ values observed at last sampling point T10. The well-defined correlation states that the comprehensive epidemiological analysis of the selected community by considering the terminal discharge point of the drain (T10).

**Table 2:** SARS-CoV-2 RNA load in weekly monitoring sewage samples.

| **Sample Code** | **E gene\*** | **N gene\*** | **ORF1ab\*** | **RNA copies/1L\*\*\*** |
| --- | --- | --- | --- | --- |
| T3 | 28.35±1.27% | 26.85±1.30% | 27.89±0 % | 23470 |
| T4 | 27.16±1.29% | 25.39±1.65% | 21.01±0.32% | 54135 |
| T5 | 28.73±1.04% | 27.54±1.82% | 28.72±0.93% | 17954 |
| T6 | 27.42±3.36% | 26.28±4.68% | 27.14±0% | 45050 |
| T7 | 28.46±2.39% | 26.58±2.67% | 21.61±1.31% | 21757 |
| T8 | 26.03±1.96% | 24.53±3.38% | 20.06±1.21% | 119391 |
| T9 | 25.41±1.50% | 23.37±3.34% | 19.56±1.33% | 184664 |
| T10 | 27.24±1.84% | 25.71±1.28% | 21.33±0.48% | 51182 |
| **Average** | **27.35±1.83%** | **25.78±2.48%** | **23.42±0.7%** | **64700** |

**\*Represent $\bar{x}$ +RSD; \*\*Average $C_T$ of E, N and ORF1ab genes; \*\*\*RNA copies (based on E gene) were calculated based on the linear fit equation.**

Table 2 depicts comprehensive data about the five-week average C_T_ values (Individual genes) of a particular sampling point, cumulative average of all points in a week along with RNA copy number. The Higher RNA copies of 184,664 RNA Copies/L were observed at sampling station T9 (Tarnaka lateral drain) where the lowest E-gene CT of 25.41±1.50% depicted. At the sampling point T7 (Errakunta lateral drain) a lower viral load of 17,954 RNA Copies/L was observed (Fig. 2; Table 2). On average 64,700 RNA Copies/L was observed in all the 8 points (Table 2). During the weekly longitudinal monitoring, there was not much difference noticed with the temporal (weekly) variation and therefore, one-day sampling in a week might provide comprehensive/representative load over the wind period of one week.

**Fig. 2:**
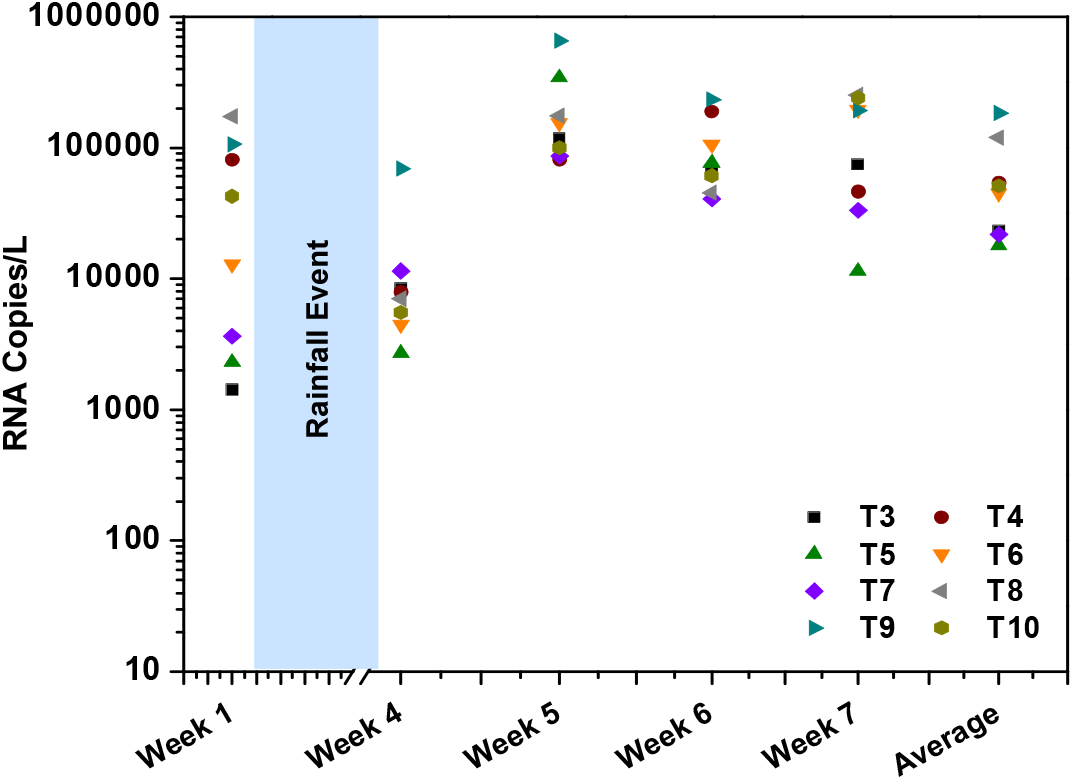
RNA copies calculated for based on linear fit equation of E-gene.

### 3.2 Monthly Sample Analysis

Apart from the weekly monitoring, the dispersive and dynamic viral presence in the domestic sewage was also assessed with long-term (monthly) analysis by selecting the final drain point (T10) as a sampling station along with some other stations (T6 to T10). Presence of the three target genes were detected in all the 7 months’ samples with variable RNA copy numbers. In July, the C_T_ values of the E-gene, N-gene, and ORF1ab were 27.38±0.36%, 26.12±1.38%, and 28.02±1.92%, respectively with 46,527 RNA Copies/L. After the first month of sampling, multiple rainfall events occurred due to the seasonal monsoon across the Deccan plateau. Because of overflow in the drains, sample collections were paused in August and September and were resumed after the flow became normal. C_T_ values of the E-gene, N-gene, and ORF1ab were observed to be 27.5±2.65%, 26.82±0.63%, and 27.34±4.10% with 42,772 RNA Copies/L which is more or less similar to July viral load analysis data. As weekly sampling was performed during October, samples collected at all selected longitudinal points were analyzed and the average values of individual genes of the eight points (T3 to T10; C_T_ values (average) 28.34±1.25 (E-gene), 27.24±1.25 (N-gene), 27.92±2.09 (ORF1ab) is reported. Specifically, the samples collected during November 2020 showed lesser C_T_ values (25.03±0.88% (E-gene), 23.26±0.73% (N-gene), 24.41±0.86% (ORF1ab) and a higher viral load (241,722 RNA Copies/L) compared to other monthly samples. This suggests the possibility of high infection rate during November 2020. In December, a considerable drop in the viral load (20,624 RNA Copies/L; C_T_ values (28.5±0.21% (E-gene), 26.84±0.73% (N-gene), 27.19±0.49% (ORF1ab)) was recorded indicating the tapering of infection within the community, suggesting a decrement in the infection rate. To maintain the accuracy of sampling, from January 2021 samples were collected in 5 sampling stations among the eight sampling stations. The viral load during January 2021 further reduced (2036 RNA Copies/L; C_T_ value-30.44±0.84% (E-gene), 29.48±4.81% (N-gene), 28.99±2.15% (ORF1ab)). However, compared to January 2021, February 2021 samples showed a marginal increment in viral load (5228 RNA Copies/L with CT of 31.10±1.98% (E-gene), 31.23±2.95% (N-gene), 28.77±2.33% (ORF1-ab)) and decreased in subsequent month of March-2021(March-2021: 2781 RNA Copies/L; C_T_ values, 31.39±2.86% (E-gene), 29.38±1.30% (N-gene), 27.63±3.38% (ORF1-ab) (Fig. 3 and Table 4). Among the six months of study, a greater number of samples were analyzed during the months of October and November because of the weekly sampling and monitoring conducted during that period.

**Table 4:**
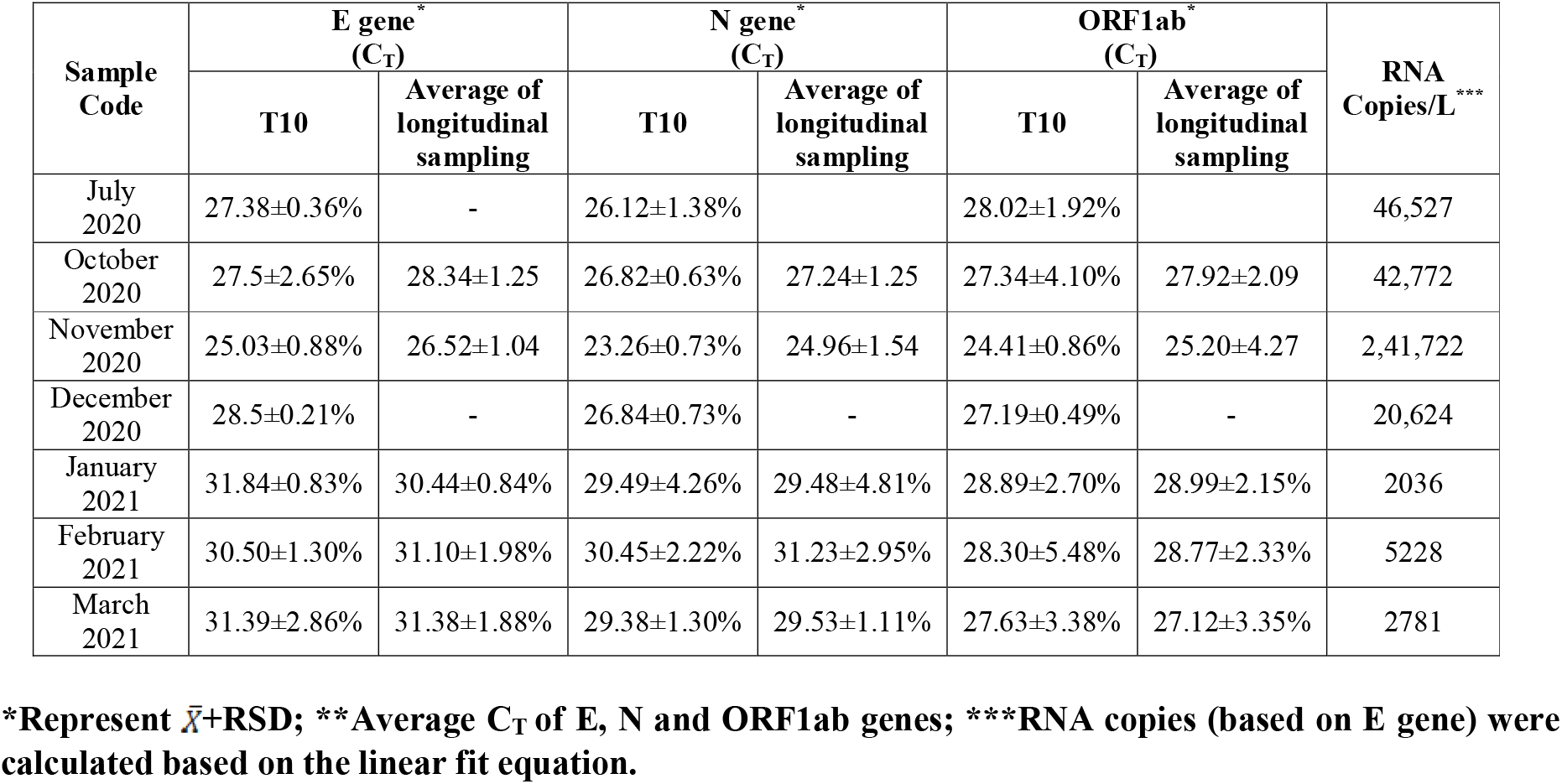
SARS-CoV-2 RNA load with monthly monitoring domestic sewage samples.

**Table 5:** Infection rate estimation through the number of people infected (symptomatic, asymptomatic, and recovered) during the sampling window.

|  |  | 10<sup>7</sup> copies/mL faeces |  |  | 10<sup>6</sup> copies/mL faeces |  |  |
| --- | --- | --- | --- | --- | --- | --- | --- |
| Sample | Capacity of the STP (MLD) | Per person contribution to STP | Method 1 | Method 2 | Per person contribution to STP | Method 1 | Method 2 |
| T3 | 18 | 67 | 330 | 352 | 6.7 | 3300 | 3520 |
| T4 |  |  | 761 | 812 |  | 7613 | 8120 |
| T5 |  |  | 252 | 269 |  | 2525 | 2693 |
| T6 |  |  | 634 | 676 |  | 6335 | 6758 |
| T7 |  |  | 306 | 326 |  | 3060 | 3264 |
| T8 |  |  | 1679 | 1791 |  | 16789 | 17909 |
| T9 |  |  | 2597 | 2770 |  | 25968 | 27700 |
| T10 |  |  | 720 | 768 |  | 7197 | 7677 |
| Estimate of infected individuals (for Study Area with 18 MLD) |  |  | 910 | 971 |  | 9098 | 9705 |
| Average estimate of infected individuals |  |  |  | 940 |  |  | 9400 |
| Estimate of the population in active phase of the infection during the window period of 77 days |  |  |  | 171 |  |  | 1709 |
| For 1800 MLD (on total Sewage Generation of Hyderabad city) |  |  | 91,000 | 97,100 |  | 9,09,800 | 9,70,500 |
| Average estimate of infected individuals |  |  |  | 94,050 |  |  | 9,40,150 |
| Estimate of the population in active phase of the infection during the window period of 77 days |  |  |  | 17,100 |  |  | 1,70,936 |

**Fig. 3:**
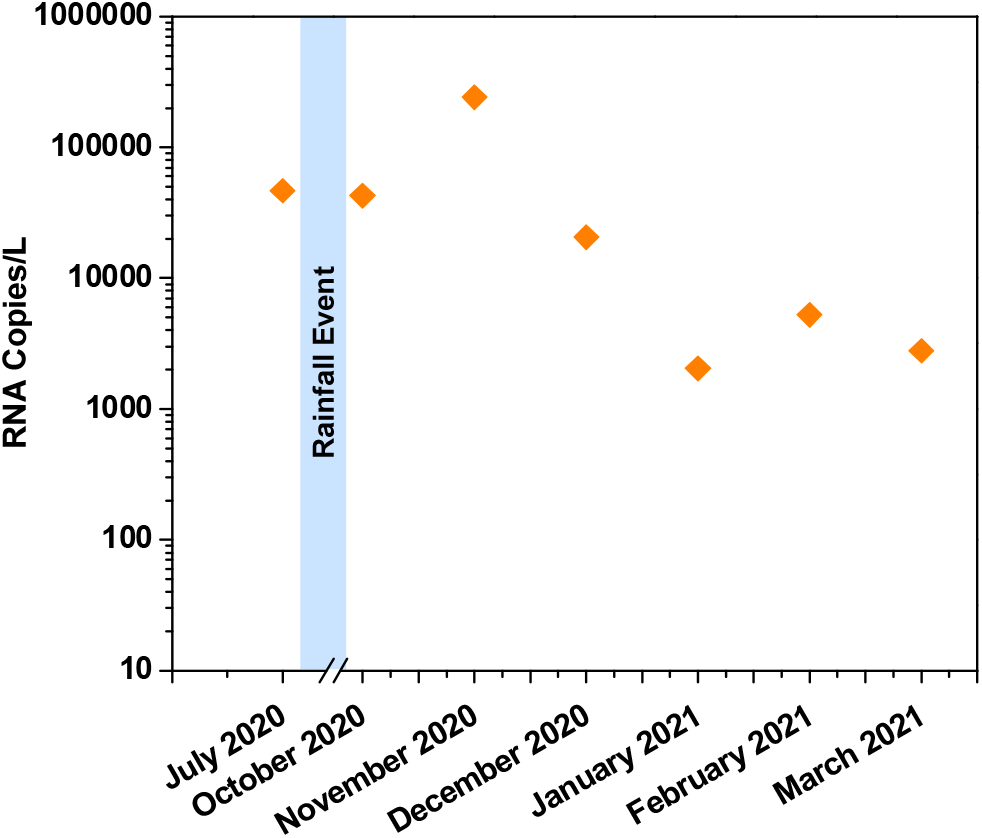
RNA copies calculated for based on linear fit equation of E-gene. The experiments were performed in triplicates.

Lower RNA copy number was observed from December till the last month of sampling (March-2021). In seven months, window period, C_T_ values of E-gene ranging from 25.03±0.88% to 31.84±0.83%. Whereas N-gene and ORF1ab ranging between 23.26±0.73% to 30.45±2.22% and 24.41±0.86% to 28.89±2.70% respectively (Fig. 3; Table 4). Temporal variation in the number of infected individuals was observed in our analysis. Such variation might be caused by various factors including infection rate, loss of viral RNA during transit from the source to the sampling site, presence of deteriorating agents in the wastewater samples, and differences in the amount of virus shed by infected individuals. Reports show a loss of 0.02 to 3000 RNA copies/mL during the passage of faecal matter from the point of defecation to the sewage drain (Foladori et al., 2020).

### 3.3 Epidemiological Analysis

Based on the RNA copy number detected in the weekly samples, enteric virus community spread was predicted by considering the volume of sewage discharge as well as the population of the selected community. Estimated numbers of infected individuals in the selected community were calculated based two methods (Ahmed et al., 2020; Hellmer et al., 2014) using two different values for the number of RNA copies shed through faeces - 10^6^ and 10^7^ RNA copies/mL faeces (Hemalatha et al., 2021; Kopperi et al., 2021). The number of infected individuals in the studied community is based on a total window period of 77 days (which includes 14 days each before and after the sampling period combined with 49 days of sampling period). The window period was selected based on the reports that showed the persistence of SARS-CoV-2 genetic material in the faecal matter of infected individuals before, during, and after the active infection phase (Wu et al., 2020; Woefel et al., 2020; Holshue et al., 2020; Peccia et al., 2020). The probable number of infected individuals in the studied community (77 days window period) was 940 with 171 people being in their active phase of infection. The community under study covers a population of ∼1.8 lakhs which represents ∼1.79% of the total Hyderabad city population. Considering the number of infected individuals in the selected community with 18 MLD domestic sewage flow (i.e. 1% of total sewage flow of 1800 MLD of Hyderabad city (https://timesofindia.indiatimes.com/city/hyderabad/master-plan-report-on-sewageby-dec-)), extrapolation was made to arrive at the total infected individuals of the city which was calculated to be 94,050 with about 17,100 active phase individuals. The infection rate of 52.2 Person/MLD was derived based on infected individuals and population figures.

Using the monthly monitoring data, the number of infected individuals was calculated during each month individually (Table 6). The number of infected individuals in the month of July was recorded as 676 (study area) and 67,606 (total city) with active phase individuals of 322 and 32195. After the rainfall event during August and September months, number of infected individuals reported in October 2020 was more or less similar to that in July 2020, wherein infected/active phase individuals in the study area and the entire city were 622/296 and 62153/26597, respectively. Marginal variation from July to October indicates the consistent spread of the enteric viral load among the community. However, in November 2020, the infection raised significantly (infected/active, 3513/1673 (study area); 351252/167263 (City)). A substantial increment in the number of infected individuals in November 2020 suggests widespread SARS-CoV-2 infection in the community. In subsequent months, December 2020 showed a substantial drop in infection (300/143 Study Area); (29970/14271 City). This decrement was observed in January 2021 with a minimal number of infection [(infected/active, 30/14 (study area); 2958/1409 (Hyderabad)]. However, February 2021 data indicated a minor increment in infected individuals of 76 and 759 in the study area and Hyderabad city, respectively, having an active phase individual of 36 (Selected community) and 3618 (Hyderabad) compared to January 2021 and followed by decreased in march-2021 (41/20 study area; 4100/1952 in city). Altogether, a higher number of infected individuals were reported in the month of November 2020 and the lowest infection was recorded in January 2021 and march-2021. The Infection rate showed more or less similar load from July to Oct with the incremental trend in Nov 2020 followed by a decrement in the next four months (Dec 2020 to Mar 2021). The repeated detection of the viral RNA over the months in sewage indicates infection severity and persistence of SARS-CoV-2.

**Table 6:** Disease dynamics and Infection rate estimation through the number of people infected (symptomatic, asymptomatic, and recovered) during the sampling window of six months.

| Sampling Month | Average estimate of infected individuals (In study area of 18 MLD) |  | Estimate of the population in active phase of the infection during the window period (29 days) |  | Estimate of infected individuals of Hyderabad city (with total Sewage generation of 1800 MLD) |  | Estimate of the population in active phase of the infection during the window period |  | Infection Rate (Person/MLD of Sewage) |  |
| --- | --- | --- | --- | --- | --- | --- | --- | --- | --- | --- |
|  | 10<sup>7</sup> copies/mL feces | 10<sup>6</sup> copies/mL feces | 10<sup>7</sup> copies/mL feces | 10<sup>6</sup> copies/mL feces | 10<sup>7</sup> copies/mL feces | 10<sup>6</sup> copies/mL feces | 10<sup>7</sup> copies/mL feces | 10<sup>6</sup> copies/mL feces | 10<sup>7</sup> copies/mL feces | 10<sup>6</sup> copies/mL feces |
| July 2020 | 676 | 6,761 | 322 | 3219 | 67,606 | 67,6092 | 32195 | 321949 | 38 | 376 |
| October 2020 | 622 | 6,215 | 296 | 2960 | 62,153 | 621,533 | 29597 | 295968 | 35 | 345 |
| November 2020 | 3,513 | 35,125 | 1673 | 16726 | 351252 | 3512,517 | 167263 | 1672672 | 195 | 1951 |
| December 2020 | 300 | 2,997 | 143 | 1427 | 29,970 | 299,699 | 14271 | 142714 | 17 | 166 |
| January 2021 | 30 | 296 | 14 | 141 | 2958 | 29581 | 1409 | 14086 | 2 | 16 |
| February 2021 | 76 | 760 | 36 | 360 | 7597 | 75954 | 3618 | 36178 | 4 | 42 |
| March 2021 | 41 | 404 | 20 | 192 | 4100 | 40400 | 1952 | 19238 | 2 | 22 |

WBE studies are effective strategies to track the disease dynamics of viral infections. Longitudinal sampling from the different selected points given an appropriate state of viral infection within the selected community (Alygizakis et al., 2020; Dhama et al., 2021). The number of infected individuals reported in the present study includes pre- and post-symptomatic, asymptomatic, and mildly symptomatic individuals. Associated with clinical data, WBE could provide critical monitoring of SARS-CoV-2 transmission within a community including the beginning, tapering, or reemergence of the virus (Gundy et al., 2009; Ahmed et al., 2020; Elsamadony et al., 2021).

The WBE studies of infectious pathogens offers unbiased monitoring of infection prevalence, spreading rate, and dynamics of infection in terms of special and temporal avenues. In conclusion, WBE studies offers an early warning system as well as provides clear view of infection dynamics and immunity status of the population, as asymptomatic, symptomatic, and pre-symptomatic individuals shed virus and the findings are unbiased. Performing WBE studies can be extended to the surveillance of other enteric infectious pathogen as the method is simple to perform yet efficient enough to help understand the infection type and dynamics among the population in a temporal manner.

## Authorship contribution statement

Athmakuri Tharak: Methodology, Investigation, Data curation, Writing – original draft. Harishankar Kopperi: Methodology, Investigation, Writing – original draft. Manupati Hemalatha: Methodology, Investigation, Data curation, Writing – original draft. Uday Kiran: Methodology, Investigation, Data curation, Writing – original draft. C.G. Gokulan: Methodology, Investigation, Formal analysis, Writing – original draft. Shivranjani Moharir: Methodology, Investigation. Rakesh K. Mishra: Conceptualization, Supervision, Funding acquisition, Validation, Writing – review & editing. and S. Venkata Mohan: Conceptualization, Methodology, Supervision, Validation, Writing – original draft, Writing – review & editing.

## Declaration of competing interest

The authors declare that they have no known competing financial interests or personal relationships that could have appeared to influence the work reported in this paper.

## Data Availability

The data of the manuscript is given in the uploaded file

## Acknowledgments

The work was supported by Council of Scientific and Industrial Research (CSIR), New Delhi, India in the form of project entitled ‘Testing for COVID-19 in wastewater as a community surveillance measure (6/1/COVID-19/2020/IMD)’. UK thanks UGC, CGG and MH thank CSIR for the financial support received. KH, AT, MH and SVM acknowledge the Director, CSIR-IICT for the support.

## Notes

### Competing Interest Statement

The authors have declared no competing interest.

